# Epidemiology and treatment trends in lumbar and cervical spinal canal stenosis beyond the COVID-19 pandemic – a nationwide analysis

**DOI:** 10.1101/2025.10.26.25338840

**Authors:** Christopher Krämer, Ahmad Almahozi, Judith Rösler, Joan Alsolivany, Christina Susanne Mark-Sargut, David Wasilewski, Nora F. Dengler, Katharina Faust, Tarik Alp Sargut

**Author notes:** Corresponding Author: Tarik Alp Sargut, MD, Department of Neurosurgery, Charité – Universitätsmedizin Berlin.

## Abstract

**Study Design:** Retrospective, population-based cohort study

**Objective:** The global incidence of degenerative spinal canal stenosis has steadily increased in recent decades. However, comprehensive epidemiological data and trends in clinical management, especially during the COVID-19 pandemic, are lacking. We aimed to assess nationwide in-hospital treatment trends for degenerative spinal canal stenosis in Germany.

**Methods:** Data from the German Federal Statistical Office were analyzed for patients hospitalized with ICD-10 codes M48.02 (cervical) and M48.06 (lumbar) between 2014 and 2023. The acquired data was processed and analyzed using statistical modeling.

**Results:** The number of patients treated for lumbar spinal canal stenosis (LSS) increased from 78,329 in 2014 to 84,507 in 2023 (p = 0.626), and for cervical spinal canal stenosis (CSS) from 12,818 to 16,590 (p = 0.003). Only the increase in CSS reached statistical significance, particularly among elderly patients after 2020. Female patients were more frequently hospitalized for both LSS and CSS. There was a trend toward shorter in-hospital stays for both groups: LSS (8.49 to 6.65 days, p < 0.001) and CSS (8.08 to 7.1 days, p < 0.001). Hypertension, diabetes, and coronary artery disease were the most common comorbidities.

**Conclusions:** The incidence of hospital-treated LSS and CSS has increased over the past decade. For elderly patients with CSS, treatment numbers rose significantly following the COVID-19 pandemic, possibly reflecting their vulnerability to reduced physical activity. Overall, shorter in-hospital stays and shifting surgical trends suggest evolving management strategies in degenerative spinal canal stenosis.

## Introduction

Spinal canal stenosis is a major cause of back pain and neurological impairment affecting over 100 million people worldwide. (1)

Lumbar spinal canal stenosis (LSS) is a degenerative condition that can lead to compression of the cauda equina or nerve roots, typically presenting with neurogenic claudication or radicular symptoms including pain and additional motor or sensory neurological deficits. Cervical spinal canal stenosis (CSS) upon progression may cause either to nerve root compression with radicular symptoms or myelopathy as a result of a central cord compression. Both conditions can lead to devastating symptoms for patients. In the past decades, the number of surgical procedures for both LSS and CSS has rapidly increased (2–4), withup to an eightfold rise in operations for LSS being reported in the US between 1979 and 1992. Recently, lumbar spinal stenosis operations were the most rapidly increasing procedures. (5) However, epidemiological data about recent years are lacking, esspecially with respect to changes duringthe COVID-19 pandemic. During the pandemic, there was a substantial decrease in diagnosis of several conditions. (6) On the other hand, a compensatory increase of incidence of the major conditions, such as myocardial infarction and cancer, has not been reported after the pandemic. (7,8)

We therefore conducted the first comprehensive analysis of the incidence of LSS and CSS since 2013 in Germany, using administrative data from a registry of all hospitals nationwide with the purpose of identifing similar epidemiological trends and changes in surgical strategies in the recent years.

## Methods

### Data Source

Case-based hospital data was obtained from the German Federal Statistical Office (Statistisches Bundesamt) between 2014 and 2023. Data extraction was conducted for hospital admissions coded with ICD-10-GM (International Classification of Diseases, 10^th^ revision, German modification) codes corresponding to cervical and lumbar spinal canal stenosis (M4800-4899). Additionally, we identified and included relevant comorbidities using ICD-10-GM codes and surgical procedures using OPS (German procedure classification) codes. Trends in surgical procedures were also evaluated by screening associated OPS codes representing the procedure coded for the main diagnosis.

The dataset was accessed on 15 February 2025 from the Research Data Center (Forschungsdatenzentrum, FDZ) of the Federal Statistical Office of Germany (DESTATIS). All data were fully anonymized and aggregated prior to access; the authors had no access to any information that could identify individual participants during or after data collection

### Data Processing

All data processing and analysis were conducted using RStudio Version 1.2.5042. Filtering, cleaning, and visualization of data were performed using base R functions and the ggplot2 package for graphical representation. The parameters treated patient numbers, duration of in-hospital stay, age distributions, and treatments were plotted across years and diagnosis groups.

For statistical analysis basic R and Python functions and the statsmodels package in Python were used for the analysis. The utilised statistical test is indicated in the figure legend. Code avaibility is upon request. Thresholds for statistical significance were set at p < 0.05.

## Results

### Epidemiological trends in the treatment of LSS and CSS

The majority of patients suffering from LSS are aged between 60 and over 80 years (comp. Table 1 and Fig. 1 c and e). For all age groups a statistically significant increase in in-hospital treatments between 2014 and 2023 was not identified (comp. Table 1, p=0.626). Further analyzing this trend, we observed a significant increase from 2014 until 2019 from 78329 to 88997 cases (p < 0.001), however after 2019, numbers decreased significantly (comp. Table S1) during the COVID-19 pandemic as described below.

**Table 1.**
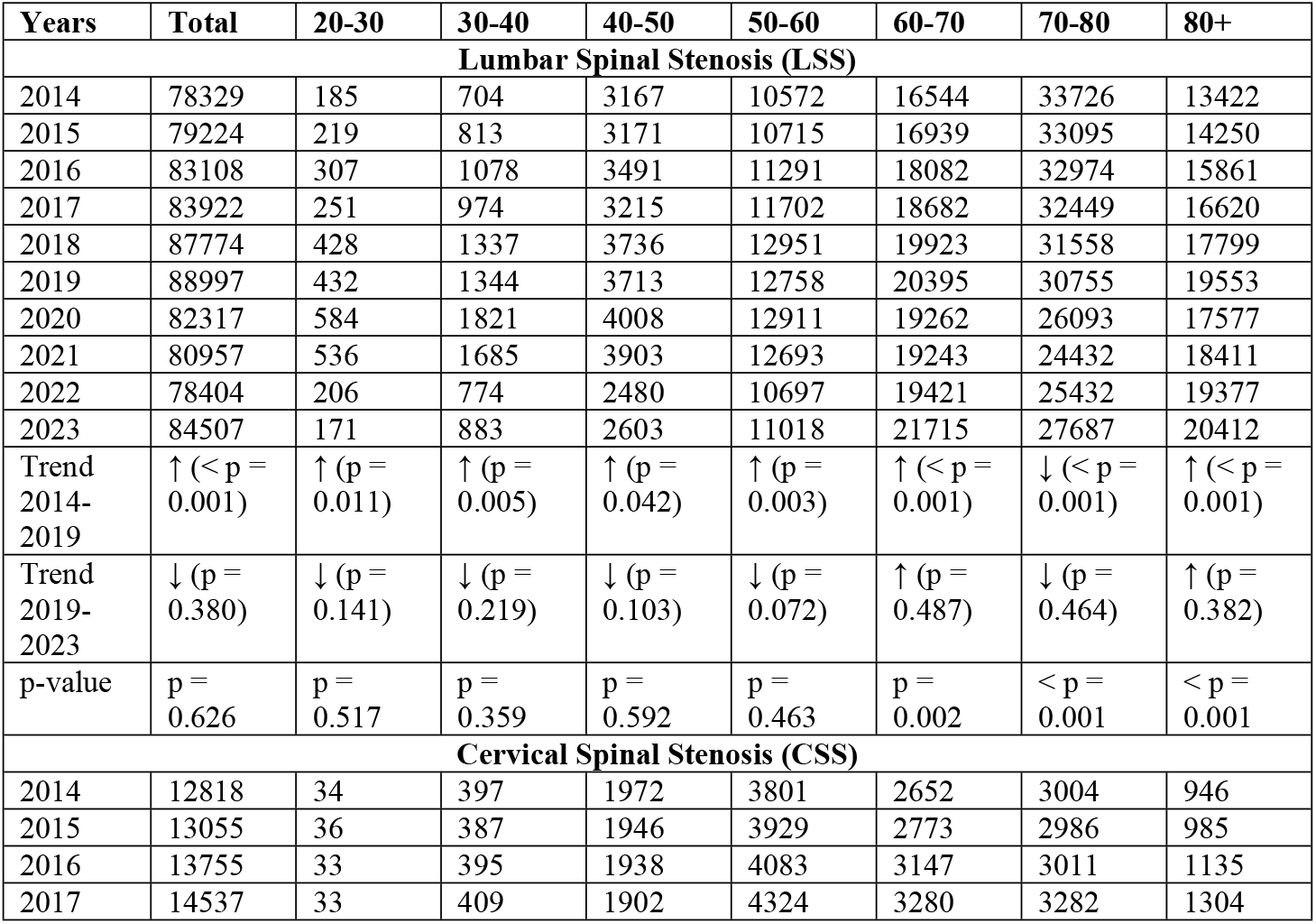
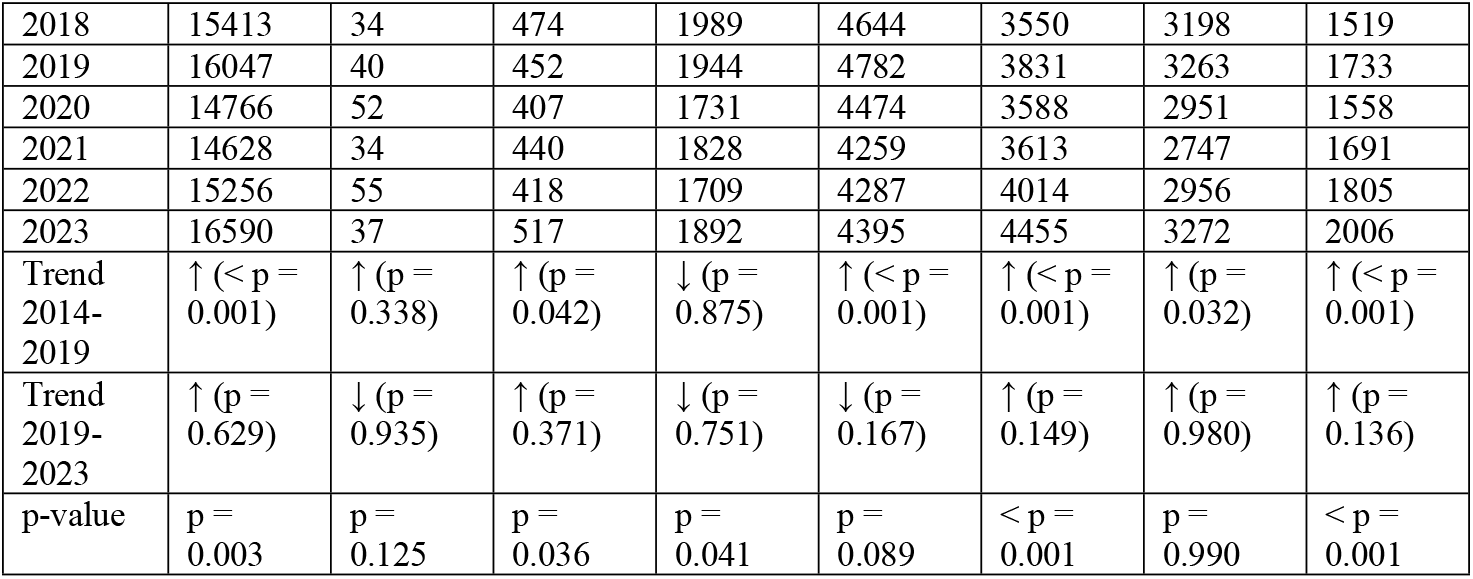
Age distribution of patients treated for LSS and CSS. Linear trend model was calculated for statistical testing (statsmodel in Python).

**Fig. 1.**
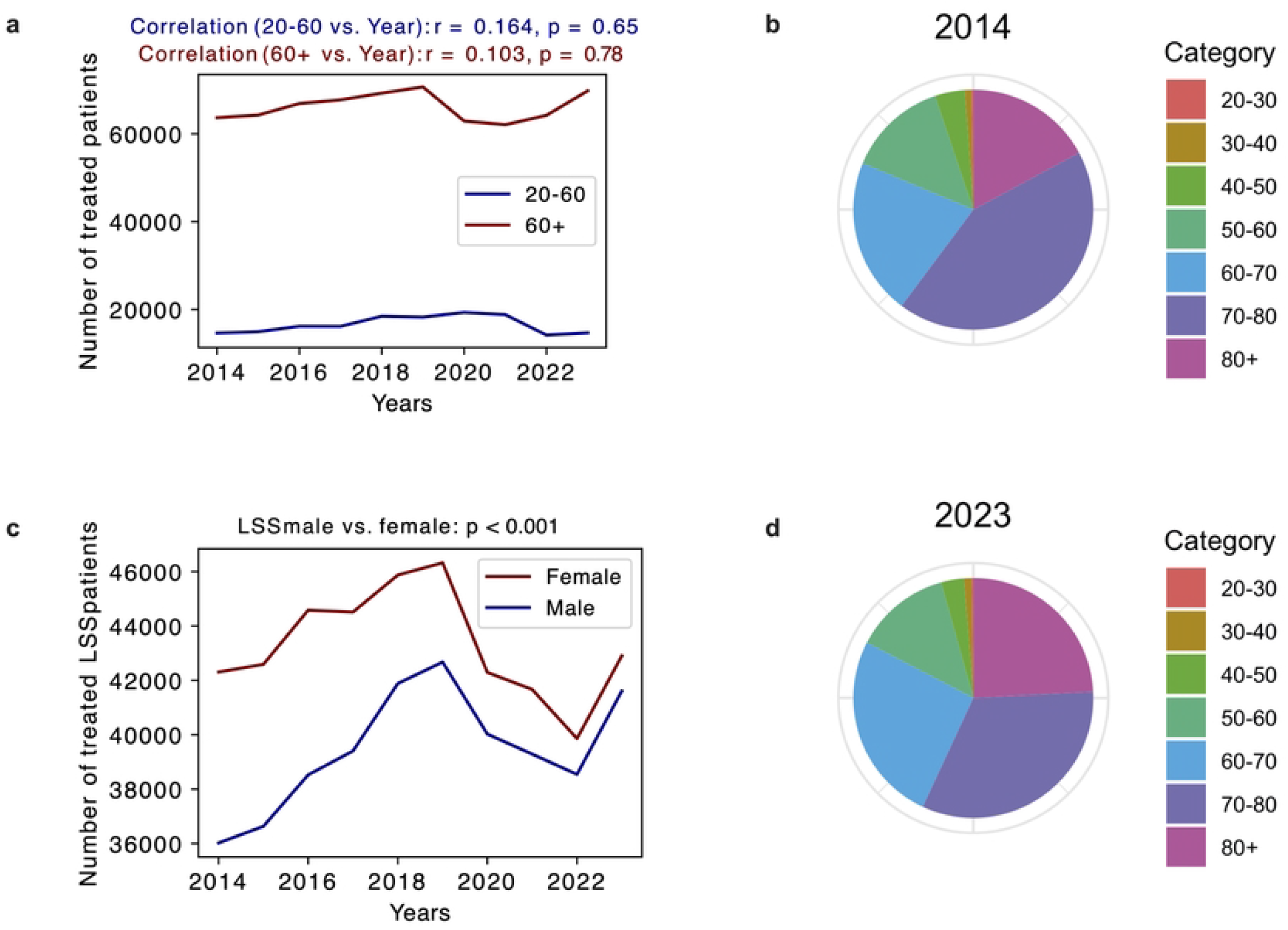
Treated patients for lumbar spinal canal stenosis between 2014 and 2023. a. Number of treated patients per year in the age groups 20-60 years (blue) and over 60 years (red). Linear correlation coefficient r and p value p for the linear regression model respectively are shown above. b. Pie chart of the age distributed number of treated patients for lumbar spinal canal stenosis in 2014. c. Gender distribution of LSS patients. Poisson regression model fitted and test results for gender female vs. male are shown. d. Pie chart of the age distributed number of treated patients for lumbar spinal canal stenosis in 2023.

Dichotomizing the population into under and over 60 years old patients, there were no significant trends over the whole time period (comp. Fig. 1b, p=6.51e-01) < 60; p=7.76e-01 > 60). Analysis of individual age groups demonstrated that patients between 60 and 70 years and over 80 years had a significant increase in treatment numbers between 2014 and 2023. However, in the group of patients aged between 70 and 80 years, a significant decrease was reported (comp. Table 1, 60-70: + 31,2 % p=0.002; 70-80: −17.9 % p < 0.001; 80+: +52.1 % p < 0.001). In the younger cohort, treatment numbers remained mostly unchanged (comp. Table 1). Over the whole time period, there were more female patients requiring in hospital treatment than males (comp. Fig. 1d, p < 0.001).

Furthermore, change in treatment numbers before and during the COVID-19 pandemic was assessed. From 2019 to 2020 all patient number over 60 years, especially in those over 70 years decreased significantly, ranging from −5.6 % to −15.2 % (comp. Table S1; 60-70: −5.6 % p < 0.001; 70-80: −15.2%, p < 0.001; 80+: −10.1%, p < 0.001). Interestingly, in younger patients we observed a relative increase in treatment numbers (comp. Table S1, 20-30: +35.2%, p < 0.001; 30-40: +35.5%, p < 0.001; 40-50: +7.9%, p= 0.001; 50-60: +1.2%, p=0.382). In contrast, after the pandemic (post 2021) the exact opposite trends are identified, with decreasing treatment numbers in younger patients (< 60 years) and increasing numbers in the older cohort between 2021 and 2022 (comp. Table S1).

For CSS, there was an overall increase in the number of patients treated in hospital between 2014 und 2023 (comp. Fig. 2a, +29.4%, p=0.003). The change in treatment numbers was driven by the over 60 age group (comp. Fig. 2b, p < 0.001). Looking into the individual groups we observed an increase of 68.0 % (mean annual increase 6.8 %) for the group 60-70 years (p < 0.001) and 112.1 % (mean annual increase 11.2 %) in patients over 80 years (p < 0.001) for the whole time period between 2014 and 2023. The age distribution demonstrates that the majority of patients were aged between 40 und over 80 years old with relatively homogenous distribution between 50 and 80 years (comp. Fig. 2a, c and e). A larger number of women required in hospital treatment compared to men (p = 0.005). Focusing on the potential effects of the COVID-19 pandemic, we observed a decrease between −11.0 % and −6.4 % for nearly all agegroups between 2019 and 2020. After the pandemic, treatment numbers,especially in the elderly population (> 60 years), rose between 6.7 % and 11.1 % in the invidiual groups between 2021, 2022 and 2023 respectively (comp. Table S2).

**Fig. 2.**
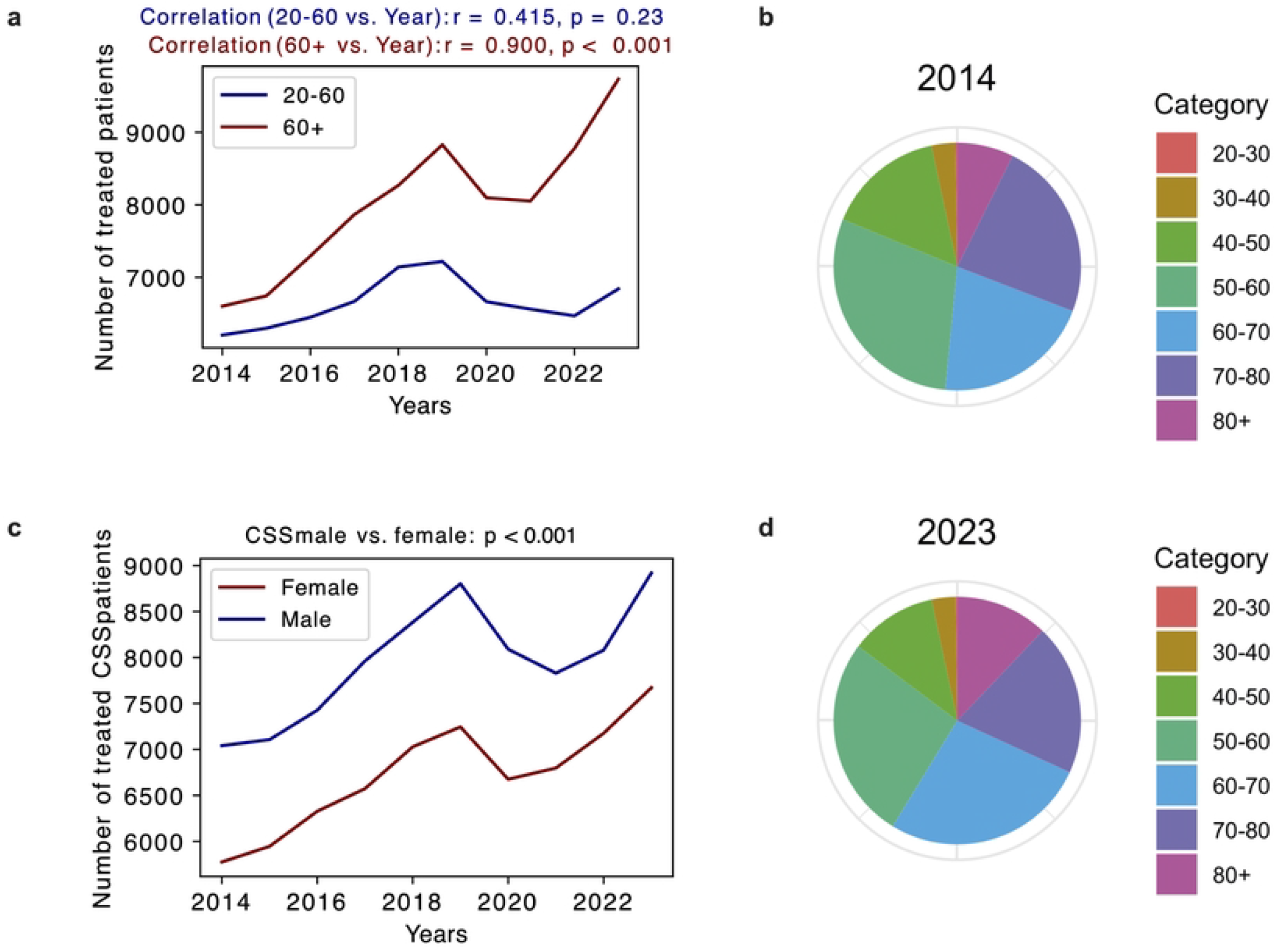
Treated patients for cervical spinal canal stenosis between 2014 and 2023. a. Number of treated patients per year in age group 20-60 years (blue) and over 60 years (red). Linear correlation coefficient r and p value p for the linear regression model respectively are shown above. c. Pie chart of the age distributed number of treated patients for cervical spinal canal stenosis in 2014. d. Gender distribution of CSS patients. Poisson regression model fitted and test results for gender female vs. male are shown. e. Pie chart

### Comorbidity burden in patients with LSS and CSS

The observed frequencies of comorbidities were similar between patients with CSS and LSS (comp. Fig. 3a and b). The most common diagnosis was arterial hypertension (comp. Fig. 3a and b). Other secondary diagnoses belonged to either metabolic disorders, such as Diabetes mellitus and fatty acid metabolism disorders or common cardiological conditions including atrial fibrillation and coronary artery disease (comp. Fig, 3a and b). There were no relevant gender dependent difference in the comorbitidies except for a higher percentage of male patients with coronary artery disease of all treated spinal canal stenosis patients (comp. Fig. 3a, b and c and Table 2).

**Table 2.** Comorbities of patients with LSS and CSS. Odds ratio and confidence intervals for male vs. female patients were calculated using statsmodel in Python.

| Comorbidity | Count male | Count female | Odds Ratio [CI] |
| --- | --- | --- | --- |
| **CSS** |  |  |  |
| Hypertension | 4270 (6.21 %) | 3256 (5.70 %) | 1.1 [1.05, 1.15] |
| DM Type 2 | 1239 (1.80 %) | 811 (1.41 %) | 1.28 [1.17, 1.39] |
| Hyperuricemia | 486 (0.71 %) | 150 (0.26 %) | 2.71 [2.25, 3.25] |
| Hypercholesterolemia | 570 (0.83 %) | 334 (0.58 %) | 1.42 [1.24, 1.63] |
| Hyperlipidemia | 750 (1.09 %) | 460 (0.81 %) | 1.36 [1.21, 1.53] |
| Atrial fibrillation | 360 (0.52 %) | 228 (0.40 %) | 1.31 [1.11, 1.55] |
| Obesity | 277 (0.40 %) | 227 (0.40 %) | 1.01 [0.85, 1.21] |
| Coronary artery disease | 881 (1.28 %) | 325 (0.57 %) | 2.27 [2.0, 2.58] |
| **LSS** |  |  |  |
| Hypertension | 24080 (8.07 %) | 24778 (7.52 %) | 1.08 [1.06, 1.1] |
| DM Type 2 | 7492 (2.51 %) | 6328 (1.92 %) | 1.32 [1.27, 1.36] |
| Hyperuricemia | 3205 (1.07 %) | 1424 (0.43 %) | 2.5 [2.35, 2.66] |
| Hypercholesterolemia | 3097 (1.04 %) | 2600 (0.79 %) | 1.32 [1.25, 1.39] |
| Hyperlipidemia | 4624 (1.55 %) | 3795 (1.15 %) | 1.35 [1.29, 1.41] |
| Atrial fibrillation | 2251 (0.76 %) | 1974 (0.60 %) | 1.26 [1.19, 1.34] |
| Obesity | 1672 (0.56 %) | 1546 (0.47 %) | 1.2 [1.12, 1.28] |
| Coronary artery disease | 6043 (2.02%) | 2638 (0.80 %) | 2.56 [2.45, 2.68] |

**Fig. 3.**
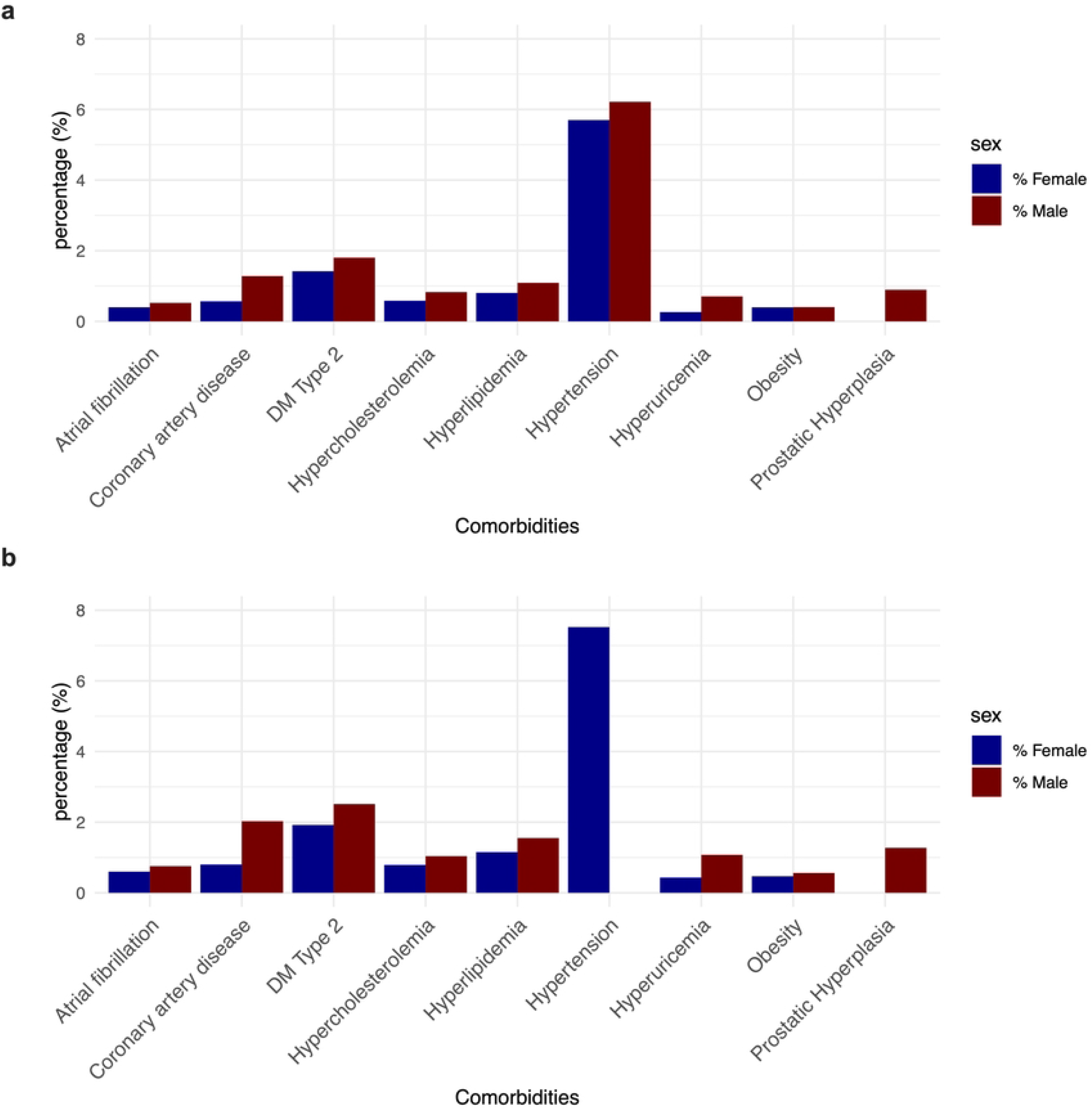
Leading comorbidities for in-hospital treated patients with cervical or lumbar spinal canal stenosis. a. Overview of comorbidities of patients treated in-hospital for cervical spinal canal stenosis distributed to age. Comorbidities were identified based on the corresponding ICD-10 code. b. Overview of comorbidities of patients treated in-hospital for lumbar spinal canal stenosis distributed to age. Comorbidities were identified based on the corresponding ICD-10 code.

### Treatment trends and length of stay in LSS and CSS

The duration of in-hospital stay significantly decreased in both groups until 2023 (Fig. 4a and 5a)., while the total duration of stay for LSS was shorter than for CSS (6.6 days (LSS) compared 7.1 days (CSS), comp. Fig. 4a). For LSS the mean duration of stay for men was consistently shorter than women over the whole time period (weighted mean duration (male): 6.86 days vs. weighted mean duration (female): 7.99 days, p-value (Welch’s t-test) < 0.001).

**Fig. 4.**
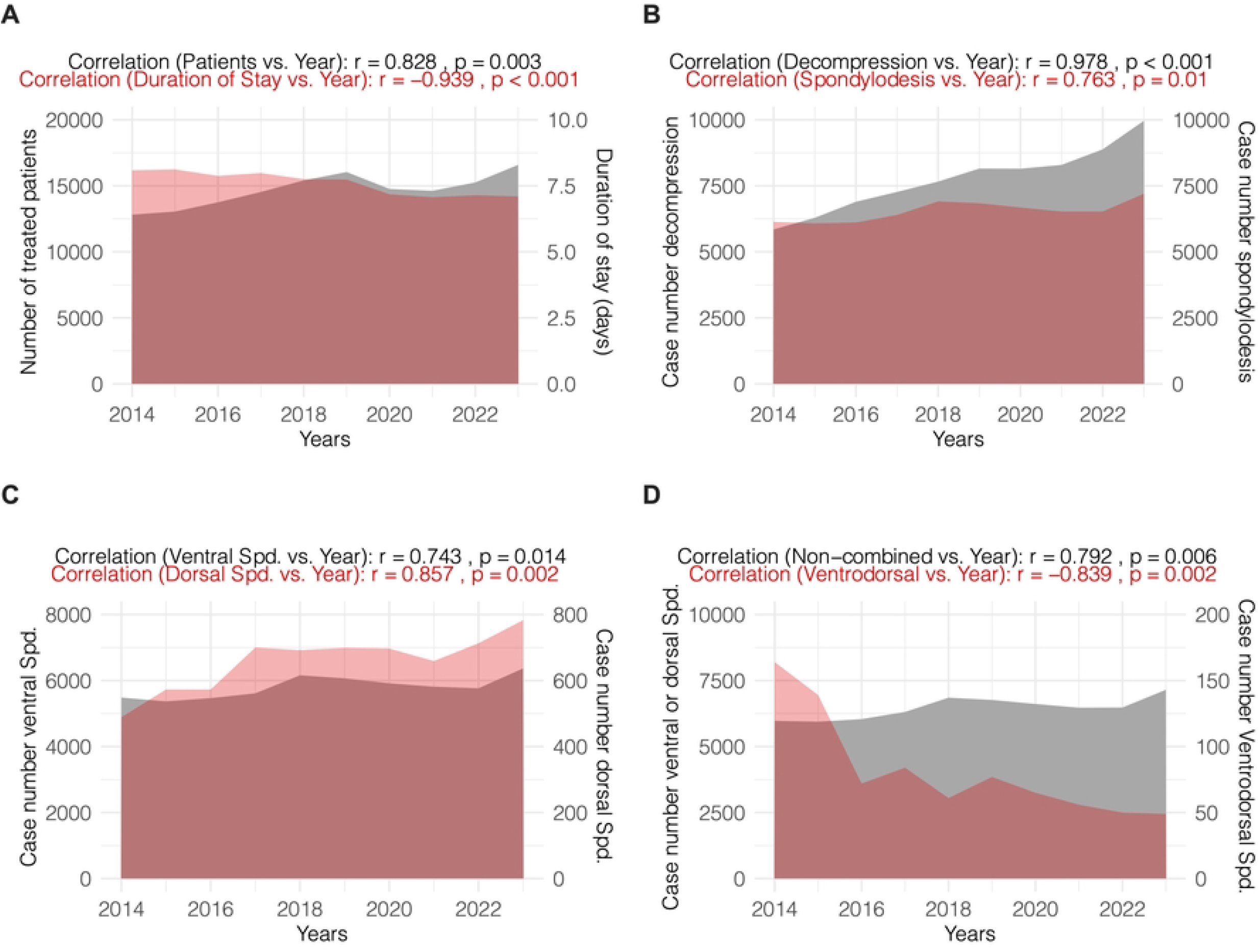
Analysis of trends in treatment of lumbar spinal canal stenosis. a. Number of patients with the diagnosis lumbar spinal canal stenosis treated with operative treatment (left y axis = colored in grey) in relation to the years 2014 – 2023. Also the changein duration of in-hospital stay in days is shown (right y axis = colored in blue). Linear correlation coefficient r and p value p for the linear regression model respectively are shown above. b. Number of patients with the diagnosis lumbar spinal canal stenosis treated with decompression without (left y axis = colored in grey) or with stabilization (right y axis = colored in blue) for the years 2014 – 2023. Linear correlation coefficient r and p value p for the linear regression model respectively are shown above. c. Number of patients with the diagnosis lumbar spinal canal stenosis treated with dorsal (left y axis = colored in grey) or ventral (right y axis = colored in blue) spondylodesis for the years 2014 – 2023. Linear correlation coefficient r and p value p for the linear regression model respectively are shown above. d. Number of patients with the diagnosis lumbar spinal canal stenosis treated with ventral or dorsal (left y axis = colored in grey) compared to combined ventrodorsal (right y axis = colored in blue) spondylodesis for the years 2014 – 2023. Linear correlation coefficient r and p value p for the linear regression model respectively are shown above.

Both decompression procedures and stabilizing approaches increased between 2014 and 2023 (comp. Fig. 4b). However, recently the numbers of anterior and lateral approaches have strongly increased compared to posterior approaches (comp. Fig. 4c). Nevertheless, looking at the total procedure counts posterior approaches were more common (comp. Fig. 4c). In the surgical treatment of LSS combined approaches using anterolateral and posterior instrumentation were more common and were performed more frequently in recent years (comp. Fig. 4d).

The duration of the hospital admission for treatment of CSS decreased significantly over time from 8.08 days to 7.1 days (comp. Fig. 5a). For CSS patients, the mean length of stay for men was longer than womenover the whole time period (weighted mean duration (male): 7.89 days vs. weighted mean duration (female): 7.23 days, p-value (Welch’s t-test) < 0.001. Procedures were split into decompressions, including partial or total laminectomy, and fusion procedures, including dorsal spondylodesis and anterior cervical discectomy and fusion (ACDF). Corresponding with the increasing number of treated patients, decompression and fusion rates both increased over time (comp. Fig. 5b). Comparing the frequency of dorsal instrumentation to ventral spondylodesis, anterior cervical discectomy and fusion was about 10 times more common (comp. Fig. 5c). Both treatment numbers increased until 2023. Lastly, comparing combined ventrodorsal procedures to the combined number of ACDF or dorsal instrumentation procedures, the ventrodorsal procedures were rarely performed and even decreased over time (comp. Fig. 5d).

**Fig. 5.**
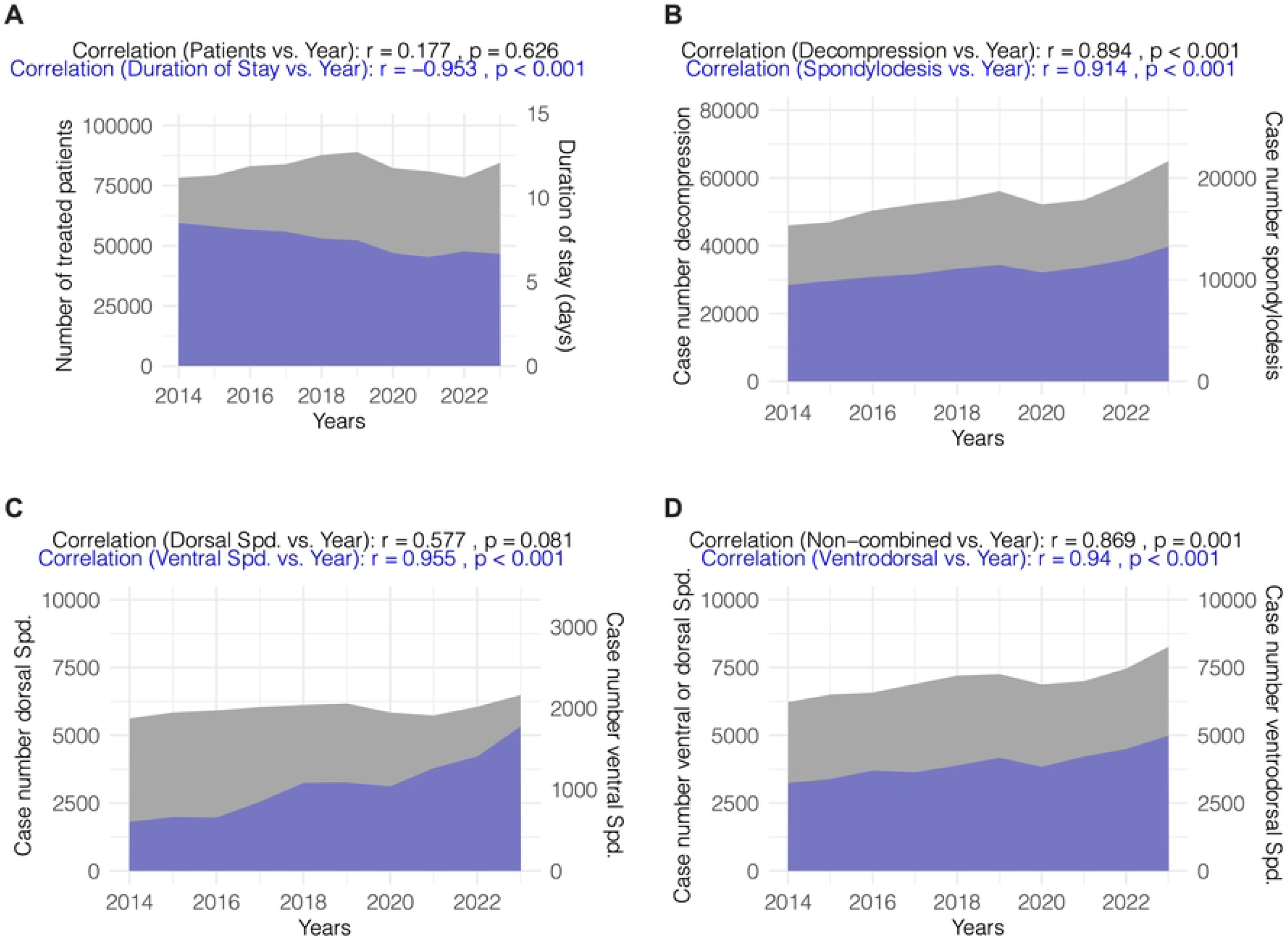
Analysis of trends in treatment of cervical spinal canal stenosis. a. Number of patients with the diagnosis cervical spinal canal stenosis treated with operative treatment (left y axis = colored in grey) in relation to the years 2014 – 2023. The trend of the in-hospital stay duration in days is also shown (right y axis = colored in red). Linear correlation coefficient r and p value p for the linear regression model respectively are shown above. b. Number of patients with the diagnosis cervical spinal canal stenosis treated with decompression either without (left y axis = colored in grey) or with stabilization (right y axis = colored in red) for the years 2014 – 2023. Linear correlation coefficient r and p value p for the linear regression model respectively are shown above. c. Number of patients with the diagnosis cervical spinal canal stenosis treated with ventral (left y axis = colored in grey) or dorsal (right y axis = colored in red) spondylodesis for the years 2014 – 2023. Linear correlation coefficient r and p value p for the linear regression model respectively are shown above. d. Number of patients with the diagnosis cervical spinal canal stenosis treated with ventral or dorsal (left y axis = colored in grey) compared to combined ventrodorsal (right y axis = colored in red) spondylodesis for the years 2014 – 2023. Linear correlation coefficient r and p value p for the linear regression model respectively are shown above.

## Discussion

This is the first study providing a comprehensive overview over the in hospital treatment of patients with cervical and lumbar spinal canal stenosis on a nationwide basis in Germany. To the author’s best knowledge, this is a novel description of the trend of case numbers of spinal canal stenosis before, during and beyond the COVID-19 pandemic. We observed a transient decrease in treatment numbers during the COVID-19 pandemic (first lockdown in Germany: 22/03/2020-06/05/2020, second lockdown: 13/12/2020-25/01/2021), which is in line with already published data from other continents. (9,10) This corresponds with a fall in surgical treatments of other degenerative conditions. (11) While treatments for lumbar spinal canal stenosis largely remained unchanged, we observed that the numbersof older patients treated for cervical spinal canal stenosis increased after the COVID-19 pandemic. Interestingly, for other diseases and also LSS in our study, a return to pre-COVID-19 levels has been observed frequently. (7,8) However, we also demostrate that reduced diagnosis and treatment of elderly patients with CSS during 2019 and 2020 resulted in a rebound of case numbers exceeding pre-pandemic case numbers. As postponed treatment of CSS is known to reduce neurological recovery(12), many elderly patients with CSS might be persistently affected by the fewer treatments during COVID-19.

We observed a significant gender difference with higher treatment numbers of women for both CSS and LSS, however, according to the current literature, the prevalence of LSS does not differ between men and women. (13) For CSS, a higher prevalence of narrowing of the spinal canal has been shown(14), which may explain the higher numbers of treated women with CSS.

Furthermore, we provide a detailed overview over surgical technique trends on a nationwide basis showing that, there is a trend towards procedures favoring shorter hospital stays. Despite introduction of the DRG system in Germany in 2003 that generally favors shorter hospital stays, there is no overalltrend towards shorter hospital length of stay in Germany. (15) For LSS, we described a stronger increase in the utilisation of procedures featuring shorter hospital stays and fewer complications in line with the existing evidence. (16) Interestingly, in other countries like the USA, other studies observed a tendency towards more complex procedures. (17) There is strong evidence that decompression surgery alone is not inferior to spondylodesis in certain conditions (18); however, in Germany, we did not observe a shift towards more decompression procedures away from lumbar fusion operations between 2014 and 2023 in all patients with LSS.

The consistent treatment numbers in older patients for LSS and the increase for CSS match the evidence that,at least for CSS, there is still a benefit in functional outcome despite comorbidities. (19)

This study has several limitations. Firstly, the data retrieved from a nationwide database did not allow individual case-based assessment of procedures, comorbidities, individual complications and clinical courses. Thus, the conclusions are only based on overall trends and therefore limited. However, this approach allows for trend analysis that is not affected by the standard procedures performed in individual hospitalsMoreover, we only assessed patient data from Germany, which limits the external validity, when comparing these numbers to other countries with different health care systems. Furthermore, as ICD-codes are based on in-hospital treatments, this study cannot describe the general epidemiology of spinal canal stenosis; here, imaging studies on healthy patients have provided estimates. (13)

## Conclusions

In summary, this study provides a detailed overview of total numbers and treatment trends for patients with lumbar or cervical spinal canal stenosis, highlighting the potential impacts of COVID-19 pandemic, on elderly patients with CSS in particular.

## Data Availability

The data underlying the results presented in the study are available from the German Federal Statistical Office (Statistisches Bundesamt, Destatis). Due to legal restrictions on the sharing of individual-level data under German data protection law, these data cannot be made publicly available. Researchers who meet the criteria for access to confidential microdata can obtain the data directly from Destatis upon formal application (contact via https://www.destatis.de).

## Acknowledgments

The authors thank the German Federal Statistical Office for providing access to the anonymized national hospital dataset used in this analysis.

## Supplemental files

**Table S1: Comparisons of yearly treatment numbers of LSS**.

Poisson regression modeling. For each year and age group the probability that the observed values differs from the expected growth rate was calculated and adjusted for multiple testing using false discovery rate (FDR) correction.

**Table S2: Comparisons of yearly treatment numbers of CSS**.

Poisson regression modeling. For each year and age group the probability that the observed values differs from the expected growth rate was calculated and adjusted for multiple testing using false discovery rate (FDR) correction.

## Notes

### Competing Interest Statement

The authors have declared no competing interest.

### Funding Statement

The author(s) received no specific funding for this work.

### Author Declarations

The present study is based exclusively on fully anonymized secondary data provided by the German Federal Statistical Office (Destatis). According to German data protection and research regulations, analyses of anonymized aggregate data do not require institutional review board (IRB) approval or informed consent. Therefore, formal ethical approval was waived.

